# Clinical Observation of Diminished Bone Quality and Quantity through Longitudinal HR-pQCT-derived Remodeling and Mechanoregulation

**DOI:** 10.1101/2022.05.11.22274961

**Authors:** Caitlyn J. Collins, Penny Atkins, Nicholas Ohs, Michael Blauth, Kurt Lippuner, Ralph Müller

## Abstract

Current clinical methods used to evaluate bone quality and quantity are insufficient for clinical evaluation of microstructural bone health, which is relevant in early diagnosis of bone disease. High resolution peripheral quantitative computed tomography (HR-pQCT) has recently emerged as a potential clinical tool for quantifying volumetric bone mineral density and microarchitecture. When combined with a longitudinal imaging protocol and finite element analysis, HR-pQCT can be used to assess bone remodeling and mechanoregulation at the tissue level. Herein, 25 patients with a contralateral distal radius fracture were imaged with HR-pQCT at baseline and 9-12 months follow-up: 16 patients were prescribed Calcium and/or Vitamin D supplement with indication of diminishing (n=9) or poor (n=7) bone quantity and 9 were not. To evaluate the sensitivity of this imaging protocol to microstructural changes, HR-pQCT images were registered for quantification of bone remodeling and image-based micro-finite element (micro-FE) analysis was then used to predict local bone strains and derive rules for bone mechanoregulation. Remodeling was predicted by both trabecular and cortical thickness and bone mineral density (R^2^>0.8), whereas mechanoregulation was affected by dominance of the arm and group classification (p<0.05). Overall, longitudinal HR-pQCT proved value in long-term patient evaluation and management of bone health.

## Introduction

Approximately 10% of older adults suffer from osteoporosis and another 40% of the same population is affected by osteopenia^1,2^. Both conditions are characterized by low bone mass and a high risk of debilitating and often life-threatening fractures. In fact, the lifetime probability of a major osteoporotic fracture caused by poor bone health (i.e. hip, spine, proximal humerus, or distal radius) is 20% in men and 50% in women^3,4^. However, patients with osteopenia are often left undiagnosed and untreated due to a more subtle deterioration of bone quality and quantity^5,6^, leaving them susceptible to further bone degeneration. When recognized clinically, patients with low bone mass are often initially recommended supplements, such as Vitamin D or Calcium. If bone mass is not increased or sustained, patients may be prescribed anti-resorptives; however, these treatments are not always effective and often have diminishing efficacy with time resulting in poor outcomes long-term.

Aside from the issues with potential treatments, a major barrier in helping patients with either osteoporosis or osteopenia is the lack of preventative screening, where fracture is often the first clinically recognized sign of low bone mass. In particular, distal radius fractures resulting from low-energy trauma are often an early warning sign for subsequent fractures at the hip or spine. By the time any of these fractures occur, patients have likely already lost large amounts of bone mass. Traditionally, bone mass is measured clinically through quantification of bone mineral content (BMC, in grams) and areal bone mineral density (BMD, in g/cm^2^) using dual-energy X-ray absorptiometry (DXA). Due to the prevalence of fractures of the radius, hip, and spine, these locations are most commonly targeted for the measurement of BMD. With the creation of larger normative and longitudinally measured databases, the use of DXA and BMD measurements has become the standard of care in the clinical diagnosis and management of osteoporosis and osteopenia. Here, diagnostic thresholds, established as standard deviations above or below a young adult reference mean (T-scores), are used to categorize patients into descriptive categories: normal (T-score ≥ −1 SD), low bone mass or osteopenia (T-score < −1 and > −2.5 SD), and osteoporosis (T-score ≤ −2.5 SD)^7^. However, measurements of BMD by DXA may not be prescribed until after a fracture^8^. In fact, the diagnosis of severe or established osteoporosis is based on both an osteoporotic T-score and one or more past fractures due to osteoporosis. Unfortunately, BMD measurements from DXA are limited to an assessment of bone quantity and have been shown to lack the necessary sensitivity to serve as an effective fracture risk assessment tool ^9–11^. In combination with individual patient risk factors in the Fracture Risk Assessment Tool (FRAX®) only marginal gains in identifying those at greater risk of fracture who would be eligible for preventative treatment have been obtained^12,13^. The lack of sensitivity of DXA, with or without the FRAX supplement, suggests that bone quality and microarchitecture play a key role in the prediction of individual fracture risk.

To better account for more subtle changes in bone quality and strength, indicative of the onset of osteopenia or osteoporosis, alternative image-based metrics have been introduced to assess bone health and fracture risk. Trabecular Bone Score (TBS)^14^, a DXA-based tool for approximating bone microstructure using texture-based analysis, has been shown to increase the prognostic value of BMD and FRAX^15,16^. However, this technique only provides a 2D assessment of the lumbar spine and has been shown to have lower reproducibility than DXA^14^. Biomechanical computed tomography (CT) analysis (BCT), an image-based finite element (FE) analysis designed to measure bone strength from clinical CT images of the hip or spine, has recently been approved for osteoporosis diagnostic testing in the United States^17^. Taking into account both 3D patient bone geometry and material properties, BCT has been used to successfully group patients into low and high fracture risk categories and provide a comprehensive measure of bone strength^17,18^. Although BCT seems to be a powerful tool for assessing bone health, varied implementations across groups and institutions result in different bone strength predictions. Due to this variability, the assessment of longitudinal changes using BCT is more robust than absolute measures^19^. However, the widespread use of longitudinal BCT is unlikely since the relatively high radiation dose of each scan (286-506 µSV for a low-dose hip CT) limits BCT to the use of scans acquired for unrelated clinical purposes^20^.

High-resolution peripheral quantitative CT (HR-pQCT), an emerging diagnostic imaging technology with low effective radiation dose (3-5 µSv), enables the assessment of 3D bone morphometrics and densitometrics, including volumetric BMD^21^, at peripheral sites such as the distal radius and tibia. In addition to direct measurement of BMD in 3D, these high-resolution images can be used to evaluate both compartment specific (i.e. cortical and trabecular) structural properties and bone mechanical properties, such as stiffness and strength through FE analysis^22,23^. A study including international patient cohorts found HR-pQCT-based estimated failure load at the tibia and radius to be the strongest predictors of incident fracture, independent of femoral neck DXA-based BMD and FRAX^6^. Further, the ability to track changes in the cortical and trabecular compartments has revealed both age-related^24–26^ and disease-specific characteristics^26–29^ not previously realized using regional DXA-based measures. However, the regional and tissue-level measures do not fully capitalize on the capabilities of HR-pQCT, which when combined with a longitudinal imaging protocol include microstructural analysis of bone remodeling^30^ and mechanoregulation^31,32^. To date, the ability of such longitudinal, extended HR-pQCT analysis tools to detect clinically relevant changes in bone quality and quantity has yet to be thoroughly investigated.

The purpose of this study was to investigate the application of longitudinal HR-pQCT imaging and associated remodeling and mechanoregulation analyses in the radius of human subjects. We hypothesized that these longitudinal, extended analyses would provide increased sensitivity to the assessment bone quality and quantity relative to traditional clinical methods. For this analysis, we utilized longitudinal HR-pQCT images of patients in three groups, those with normal bone mass and those with low bone mass, who were either prescribed no supplements or Vitamin D and/or Calcium supplements, to understand whether high-resolution 3D imaging would be useful in the clinical diagnosis and long-term management of bone health. These methods could provide the means to more accurately assess changes in bone microarchitecture for patients at risk of osteopenia and osteoporosis, overcoming the current limitations of existing clinical assessment techniques.

## Results

Of the 25 subjects, nine were not prescribed any form of additional treatment (NoSupp) while 16 were prescribed supplements (i.e., Vitamin D, Calcium, or a combination of the two), based on low values of blood-based bone markers at baseline. Of the 16 subjects who were prescribed some form of supplement, nine had no femur, lumbar spine, or radius T-Scores below −2.5 (LowSupp) and seven had at least one of these T-Scores less than −2.5 (OPSupp) (Table 1). After adjusting for pre-intervention values, neither imaging interval nor age had a statistically significant effect on post-intervention values. Therefore, these were excluded from any further analysis.

**Table 1.**
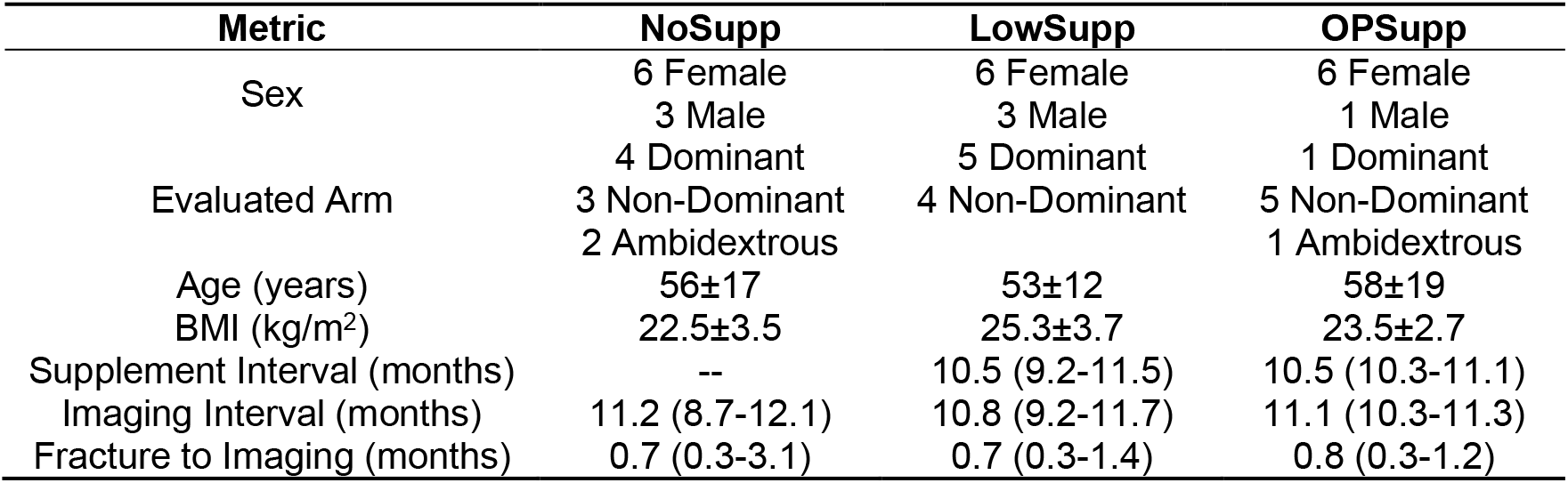
Patient demographics for subject groups. Supplement interval represents the duration of supplementation prior to the follow-up imaging session. Data presented as either mean ± standard deviation or median (range). BMI, body mass index.

### Static Morphometry and Densitometry

Subjects from the three groups had varied radius and lumbar spine T-scores as well as cortical bone mineral density (Ct.BMD) values but did not differ in other measures of bone quality or quantity (Table 2). Group, dominance of the evaluated arm, and sex were investigated covariates in the ANCOVA. Group had a near significant impact on post-intervention adjusted trabecular thickness (Tb.Th), with a magnitude of change of 2.3% (Table 3). Here, NoSupp (0.223±0.001) had significantly higher adjusted Tb.Th than LowSupp (0.218±0.002, p=0.038) but not OPSupp (0.221±0.002, p=0.68). Similarly, sex had a significant impact on adjusted cortical thickness (Ct.Th), with a magnitude of change of 15.5% (Table 3), where males had significantly higher Ct.Th (0.946±0.048) in comparison to females (0.810±0.034, p=0.025). Dominance of the evaluated arm had no significant impact on post-intervention values for any of the static morphometric parameters.

**Table 2.**
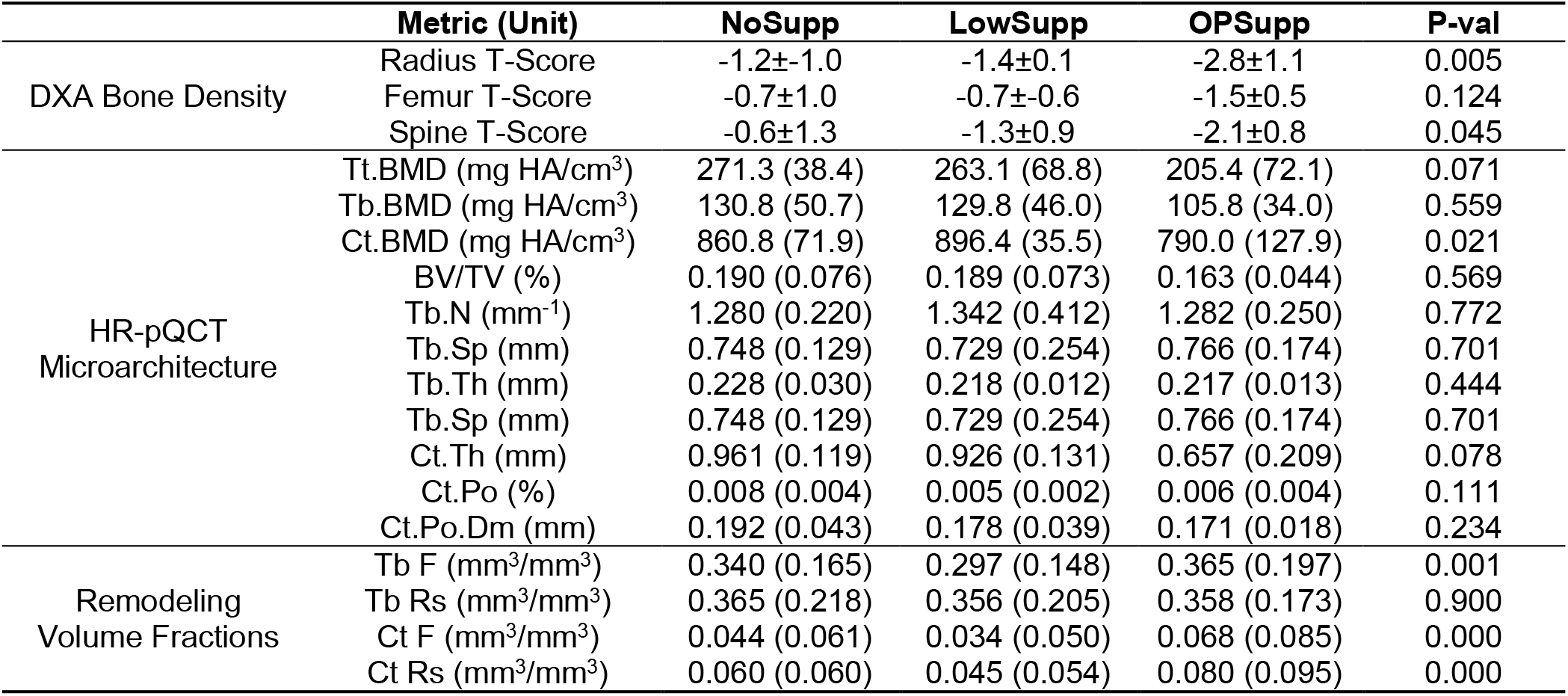
Averaged patient measures of bone morphometrics, densitometrics, and remodeling volume fractions. Data presented as either mean ± standard deviation or median (interquartile range) and averaged between measures acquired at the two evaluated timepoints for HR-pQCT microarchitecture measures, which were acquired at baseline and follow-up. P-values represent group differences as assessed by Kruksal-Wallis H test or one-way analysis of variance. A threshold of 320 mg/cm^3^ was used for remodeling volume fractions. BMD, bone mineral density; BV/TV, bone volume fraction; Tb, trabecular; N, number; Sp, separation; Th, thickness; Ct, cortical; Po, porosity; Po.Dm, cortical pore diameter; F, formation; Rs, resorption.

**Table 3.**
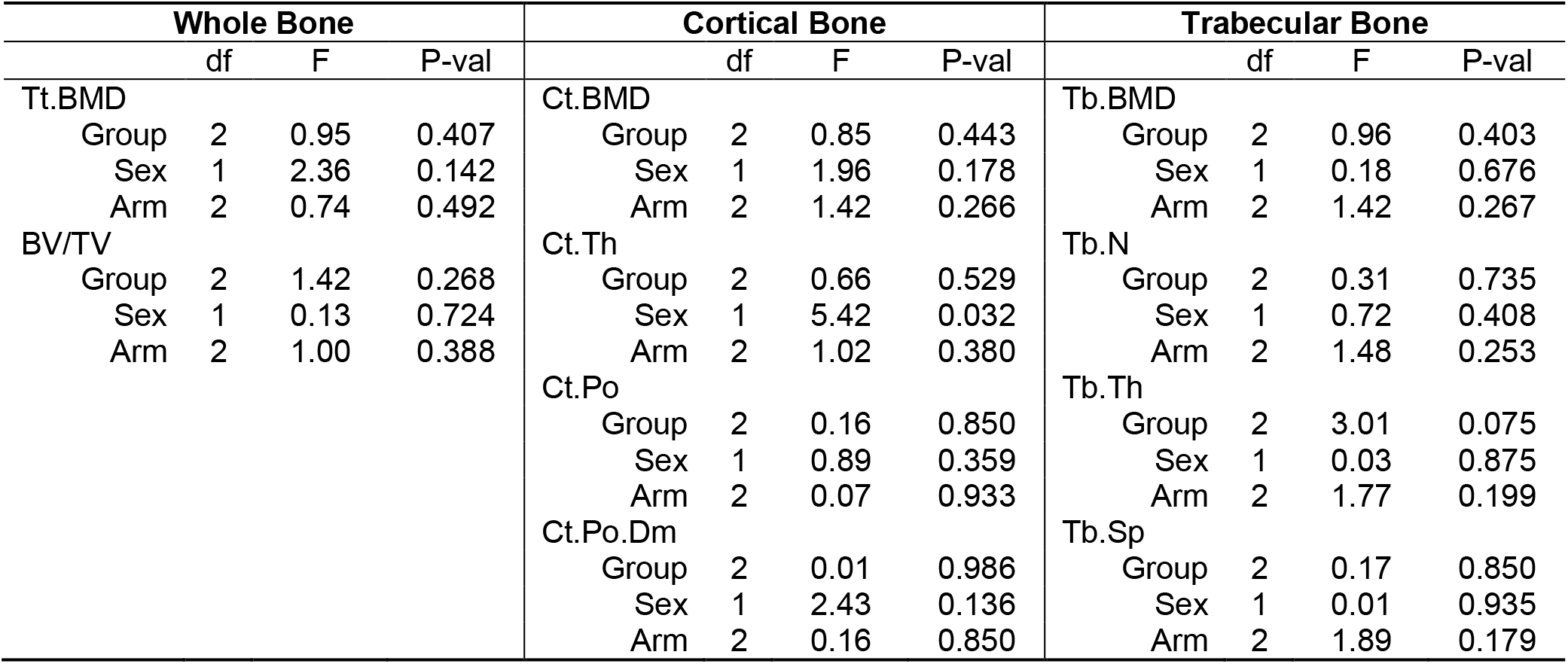
ANCOVA summary for static morphometric parameters. P-values represent contrast differences as assessed by Tukey’s HSD method. BMD, bone mineral density; BV/TV, bone volume fraction; Tb, trabecular; N, number; Sp, separation; Th, thickness; Ct, cortical; Po, porosity; Po.Dm, cortical pore diameter.

| Whole Bone |  |  |  | Cortical Bone |  |  |  | Trabecular Bone |  |  |  |
| --- | --- | --- | --- | --- | --- | --- | --- | --- | --- | --- | --- |
|  | df | F | P-val |  | df | F | P-val |  | df | F | P-val |
| Tt.BMD |  |  |  | Ct.BMD |  |  |  | Tb.BMD |  |  |  |
| Group | 2 | 0.95 | 0.407 | Group | 2 | 0.85 | 0.443 | Group | 2 | 0.96 | 0.403 |
| Sex | 1 | 2.36 | 0.142 | Sex | 1 | 1.96 | 0.178 | Sex | 1 | 0.18 | 0.676 |
| Arm | 2 | 0.74 | 0.492 | Arm | 2 | 1.42 | 0.266 | Arm | 2 | 1.42 | 0.267 |
| BV/TV |  |  |  | Ct.Th |  |  |  | Tb.N |  |  |  |
| Group | 2 | 1.42 | 0.268 | Group | 2 | 0.66 | 0.529 | Group | 2 | 0.31 | 0.735 |
| Sex | 1 | 0.13 | 0.724 | Sex | 1 | 5.42 | 0.032 | Sex | 1 | 0.72 | 0.408 |
| Arm | 2 | 1.00 | 0.388 | Arm | 2 | 1.02 | 0.380 | Arm | 2 | 1.48 | 0.253 |
|  |  |  |  | Ct.Po |  |  |  | Tb.Th |  |  |  |
|  |  |  |  | Group | 2 | 0.16 | 0.850 | Group | 2 | 3.01 | 0.075 |
|  |  |  |  | Sex | 1 | 0.89 | 0.359 | Sex | 1 | 0.03 | 0.875 |
|  |  |  |  | Arm | 2 | 0.07 | 0.933 | Arm | 2 | 1.77 | 0.199 |
|  |  |  |  | Ct.Po.Dm |  |  |  | Tb.Sp |  |  |  |
|  |  |  |  | Group | 2 | 0.01 | 0.986 | Group | 2 | 0.17 | 0.850 |
|  |  |  |  | Sex | 1 | 2.43 | 0.136 | Sex | 1 | 0.01 | 0.935 |
|  |  |  |  | Arm | 2 | 0.16 | 0.850 | Arm | 2 | 1.89 | 0.179 |

### Dynamic Morphometry

Formation and resorption volume fractions of the cortical and trabecular regions increased with mineralized bone density threshold (Figure 1). Overall differences were observed among groups for trabecular formation and cortical formation and resorption using a standardized bone mineral density threshold of 320 mg/mm^3^ (Table 2); however, these differences were not significant when evaluating the volume fractions at individual thresholds. When evaluating against demographics and the average morphometric values, cortical formation and resorption volume fractions were predicted by cortical resorption or formation, respectively, and average Ct.BMD and Ct.Th (R^2^=0.937, Q^2^=0.892 and R^2^=0.881, Q^2^=0.829, respectively).Trabecular formation volume fraction was predicted by trabecular resorption, average total bone volume fraction (BV/TV), trabecular bone mineral density (Tb.BMD), and overall time between imaging sessions (R^2^=0.797, Q^2^=0.712). Conversely, trabecular resorption volume fraction was predicted by trabecular formation, average Tb.Th, and overall time between imaging sessions (R^2^=0.822, Q^2^=0.764).

**Figure 1.**
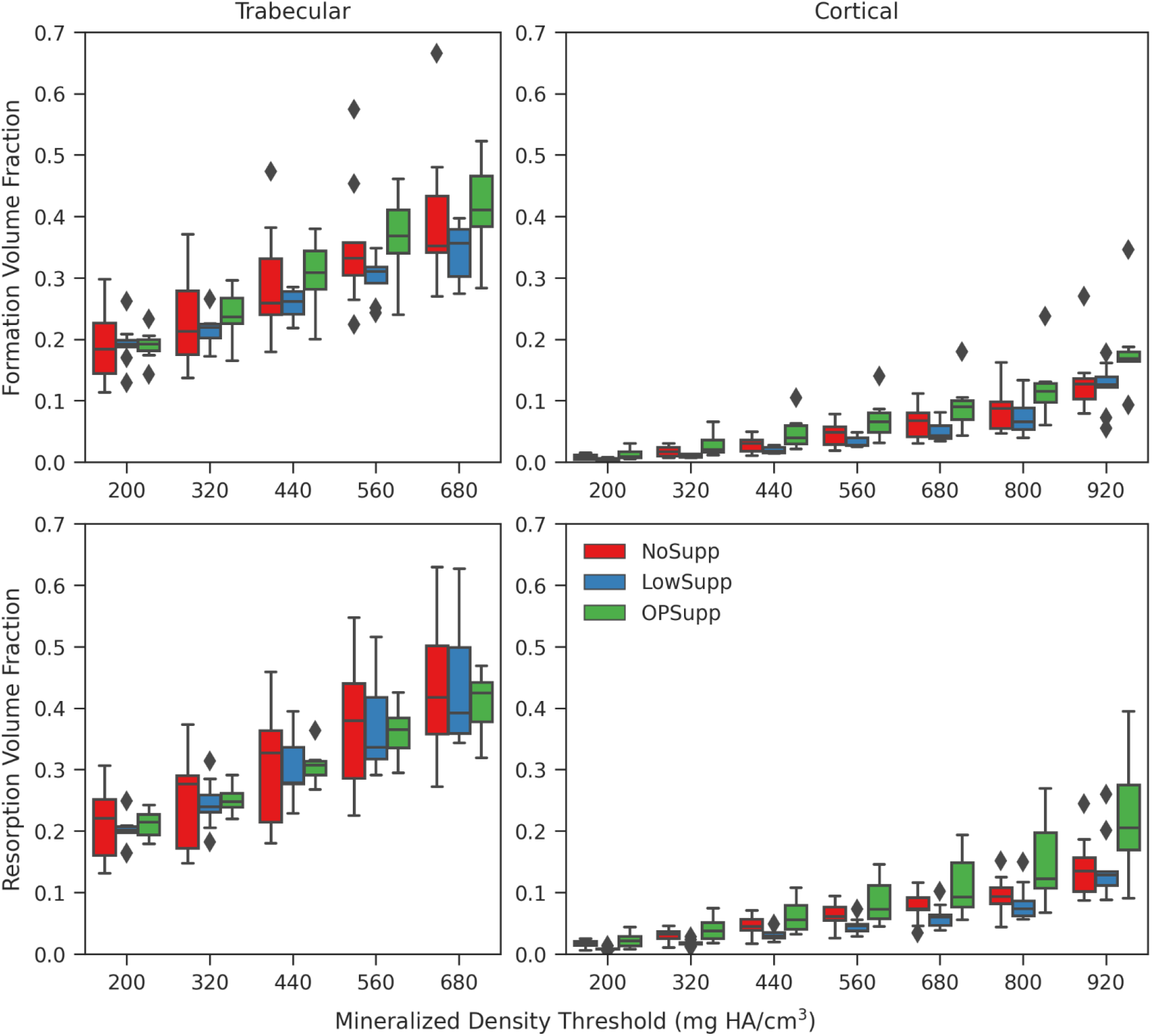
The formation and resorption volume fractions for each mineralized density threshold ranging from 200 mg Hydroxyapatite (HA)/cm^3^to 680 mg HA/cm^3^ for trabecular bone and 920 mg HA/cm^3^ for cortical bone with 120 mg HA/cm^3^ intervals.

### Mechanics and Mechanoregulation

Group, dominance of the evaluated arm, and sex were investigated as covariates in the ANCOVA. Sex had no statistically significant effect on post-intervention values for any of the measured mechanics. None of the assessed covariates had an impact on post-intervention apparent stiffness. For the cortical and trabecular effective strain measures, group and dominance of the arm had a significant (p<0.05) or near significant (p<0.10) impact on post-treatment values.

Within the cortex, group had a significant impact on 10th and 25th percentile and near significant impact on 5th percentile and median adjusted effective strain (Table 4). In comparison to LowSupp, OPSupp had significantly higher cortical adjusted effective strain at the 10th percentile (LowSupp: 2020±30 µɛ, OPSupp: 2140±40 µɛ, p=0.026), 25th percentile (LowSupp: 3050±30 µɛ, OPSupp: 3210±40 µɛ, p=0.011), and median (LowSupp: 4750±50 µɛ, OPSupp: 4940±60 µɛ, p=0.043) (Figure 2). No significant or near significant contrasts were detected among groups for the 5th or 75th percentile cortical adjusted effective strain. Arm had a significant impact on the 25th percentile and median cortical adjusted effective strain and near significant impact on 10th percentile cortical adjusted effective strain (Table 4). Ambidextrous (A) arms had significantly higher cortical adjusted effective strain than the non-dominant (ND) arms at the 10th percentile (A: 2170±50 µɛ, ND: 2020±30 µɛ, p=0.046), 25th percentile (A: 3210±40 µɛ, ND: 3050±30 µɛ, p=0.011) and median (A: 4940±60 µɛ, ND: 4750±50 µɛ, p=0.043). For the 25th percentile cortical adjusted effective strain, ambidextrous arms were also nearly significantly higher than in the dominant (D) arms (A: 3210±40 µɛ, D: 3090±30 µɛ, p=0.071).

**Table 4.**
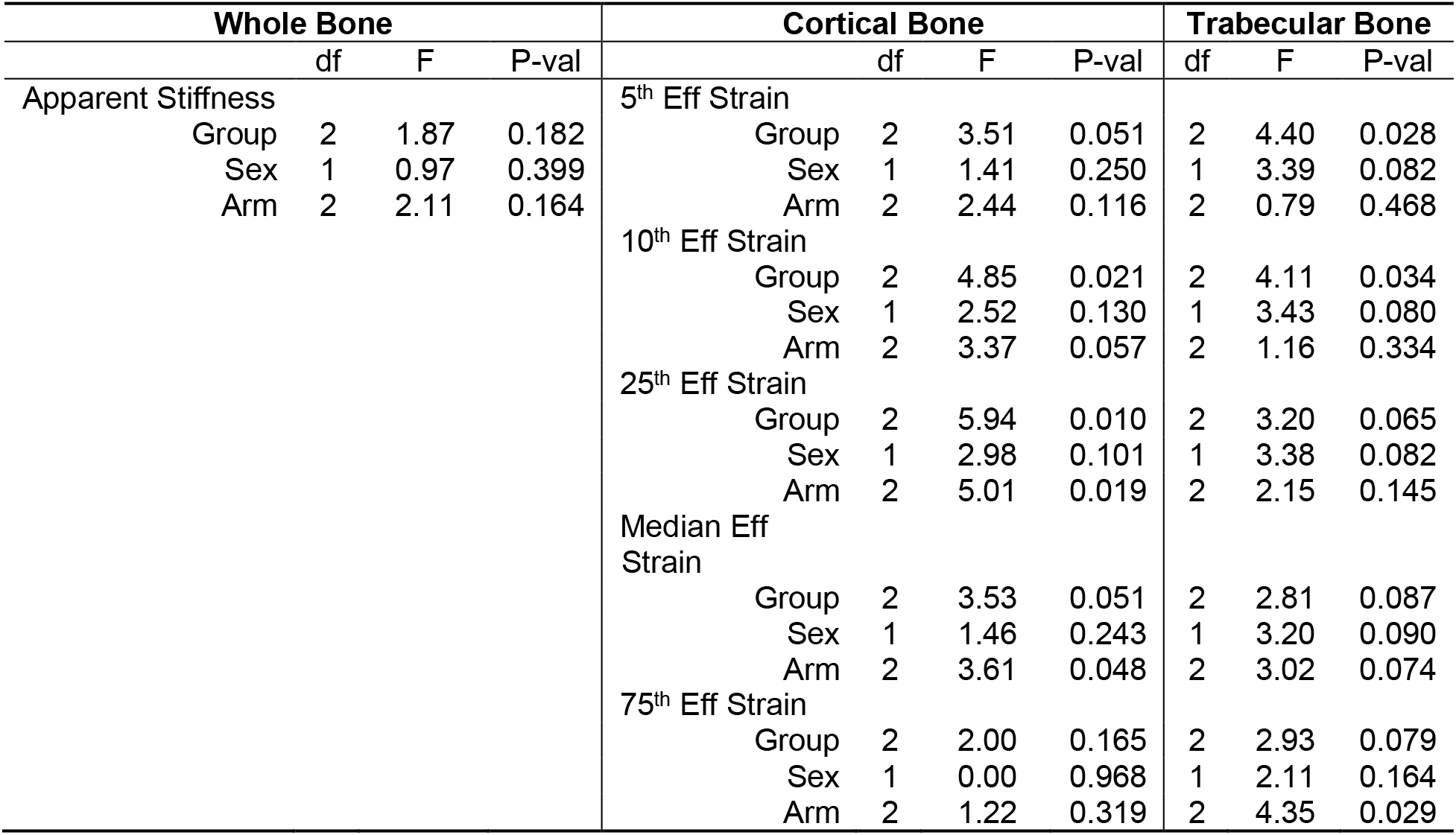
ANCOVA summary for mechanical parameters. Cortical and trabecular effective (Eff) strain values reported for 5^th^, 10^th^, 25^th^, median, and 75^th^ percentiles. P-values represent contrast differences as assessed by Tukey’s HSD method. P-values in bold are significant and those in italic are considered near significant, with significant contrasts detected during post hoc testing.

| Whole Bone |  |  |  | Cortical Bone |  |  |  | Trabecular Bone |  |  |  |
| --- | --- | --- | --- | --- | --- | --- | --- | --- | --- | --- | --- |
|  | df | F | P-val |  | df | F | P-val |  | df | F | P-val |
| Apparent Stiffness |  |  |  | 5<sup>th</sup> Eff Strain |  |  |  |  |  |  |  |
| Group | 2 | 1.87 | 0.182 | Group | 2 | 3.51 | 0.051 |  | 2 | 4.40 | 0.028 |
| Sex | 1 | 0.97 | 0.399 | Sex | 1 | 1.41 | 0.250 |  | 1 | 3.39 | 0.082 |
| Arm | 2 | 2.11 | 0.164 | Arm | 2 | 2.44 | 0.116 |  | 2 | 0.79 | 0.468 |
|  |  |  |  | 10<sup>th</sup> Eff Strain |  |  |  |  |  |  |  |
|  |  |  |  | Group | 2 | 4.85 | 0.021 |  | 2 | 4.11 | 0.034 |
|  |  |  |  | Sex | 1 | 2.52 | 0.130 |  | 1 | 3.43 | 0.080 |
|  |  |  |  | Arm | 2 | 3.37 | 0.057 |  | 2 | 1.16 | 0.334 |
|  |  |  |  | 25<sup>th</sup> Eff Strain |  |  |  |  |  |  |  |
|  |  |  |  | Group | 2 | 5.94 | 0.010 |  | 2 | 3.20 | 0.065 |
|  |  |  |  | Sex | 1 | 2.98 | 0.101 |  | 1 | 3.38 | 0.082 |
|  |  |  |  | Arm | 2 | 5.01 | 0.019 |  | 2 | 2.15 | 0.145 |
|  |  |  |  | Median Eff Strain |  |  |  |  |  |  |  |
|  |  |  |  | Group | 2 | 3.53 | 0.051 |  | 2 | 2.81 | 0.087 |
|  |  |  |  | Sex | 1 | 1.46 | 0.243 |  | 1 | 3.20 | 0.090 |
|  |  |  |  | Arm | 2 | 3.61 | 0.048 |  | 2 | 3.02 | 0.074 |
|  |  |  |  | 75<sup>th</sup> Eff Strain |  |  |  |  |  |  |  |
|  |  |  |  | Group | 2 | 2.00 | 0.165 |  | 2 | 2.93 | 0.079 |
|  |  |  |  | Sex | 1 | 0.00 | 0.968 |  | 1 | 2.11 | 0.164 |
|  |  |  |  | Arm | 2 | 1.22 | 0.319 |  | 2 | 4.35 | 0.029 |

**Figure 2.**
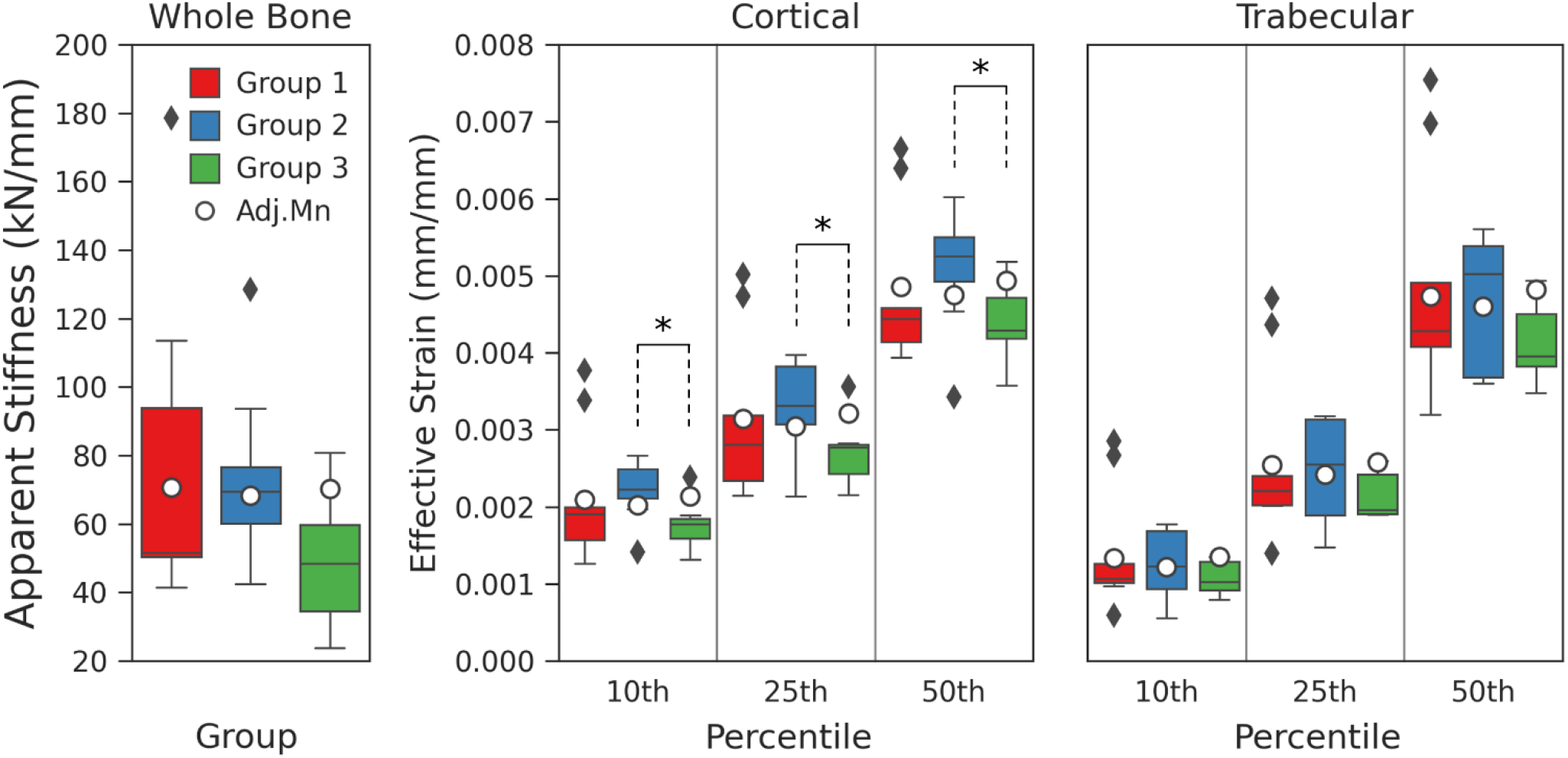
Post-treatment mechanical properties and adjusted means within the distal radius for NoSupp, LowSupp, and OPSupp. Whole bone apparent stiffness varied among groups, decreasing with treatment and worsening DXA scores; the lowest, post-treatment adjusted mean was detected in LowSupp (left). Cortical (middle) and trabecular (right) effective strain distributions, represented as discrete percentiles (10^th^, 25^th^, and 50^th^), reveal differences in post-treatment adjusted means between LowSupp and OPSupp within the cortex. (*) indicates significant contrasts between groups (p < 0.05).

Within the trabecular region, group had a significant impact on 5th and 10th percentile adjusted effective strain (Table 4, Figure 2); however, no significant differences were detected between groups in the pairwise comparison. Arm had a significant impact on 75th percentile trabecular adjusted effective strain (Table 4). Ambidextrous arms (8550±170 µɛ) had significantly higher 75th percentile adjusted effective strain than both dominant (7990±100 µɛ, p=0.025) and non-dominant (8030±80 µɛ, p=0.042) arms within the trabecular region.

The 99th percentile effective strain was not significantly different among groups (NoSupp: 28000±2300 µɛ; LowSupp: 29200±2800 µɛ; OPSupp: 26500 ± 4200 µɛ). As such, the average 99th percentile for all patients (27900 µɛ) was used to normalize the strain data from each patient for the mechanoregulation analysis. For all groups, the conditional probability (CP) of bone formation was greater at higher values of effective strain, whereas the CP of bone resorption was greater at lower values of effective strain (Figure 3). Based on these CP curves, thresholds dividing strains associated with greater probabilities of resorption and formation behavior were derived for each patient and averaged across groups. NoSupp had a lower average resorption threshold (9% strain) and higher average formation threshold (25% strain) in comparison to LowSupp and OPSupp (7% and 22% for both groups, respectively) (Figure 3). However, no significant differences in either threshold were detected among the three groups. The correct classification rate (CCR), measuring correctly classified remodeling events based on the determined resorption and formation thresholds, was similar across all groups (NoSupp = 0.408, LowSupp = 0.403, and OPSupp = 0.406) indicating consistent overall remodeling behavior. Qualitatively, regions of higher effective strain were located more distally within the analyzed region of the bone, while there were no obvious regional trends for bone remodeling (Figure 4). However, local variations in mechanics and remodeling identified that lower-level regions of higher effective strain showed increased bone quality and/or quantity over the duration of the study, i.e., formation, while regions of lower effective strain showed decreased bone quality and/or quantity, i.e., resorption (Figure 4).

**Figure 3.**
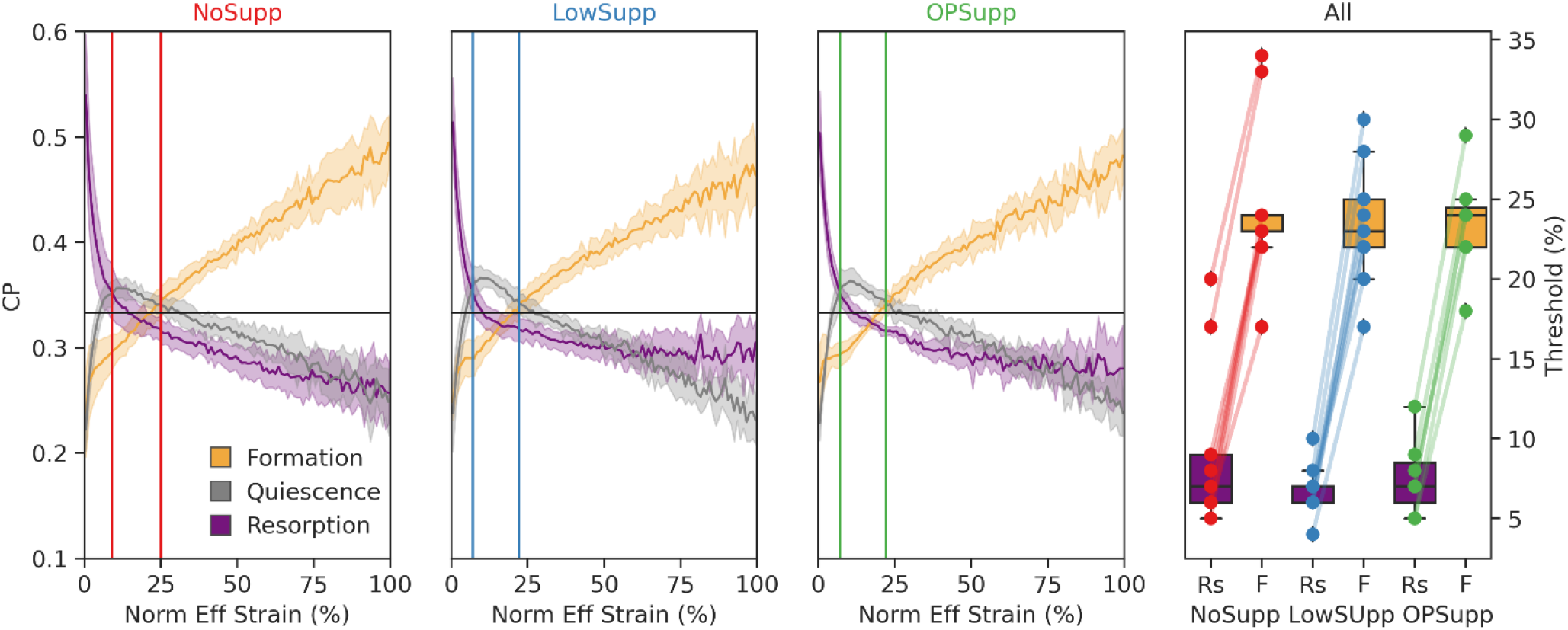
The conditional remodeling probability (CP) of remodeling sites relative to the mechanical environment, quantified as effective (Eff) strain from a simulated 1% compression, for NoSupp, LowSupp, and OPSupp groups. Normalized Eff strain distributions were used to calculate the CP for events of formation (shown in orange), quiescence (shown in grey), and resorption (shown in purple) to occur at distinct strain levels. Average thresholds dividing strains associated with resorption dominant (Rs) and formation dominant (F) probabilities each group are indicated by the left and right vertical lines, respectively, for NoSupp, LowSupp, and OPSupp (left three plots). Group and patient specific Rs and F thresholds confirmed links between bone formation at high and resorption at low mechanical signals (right).

**Figure 4.**
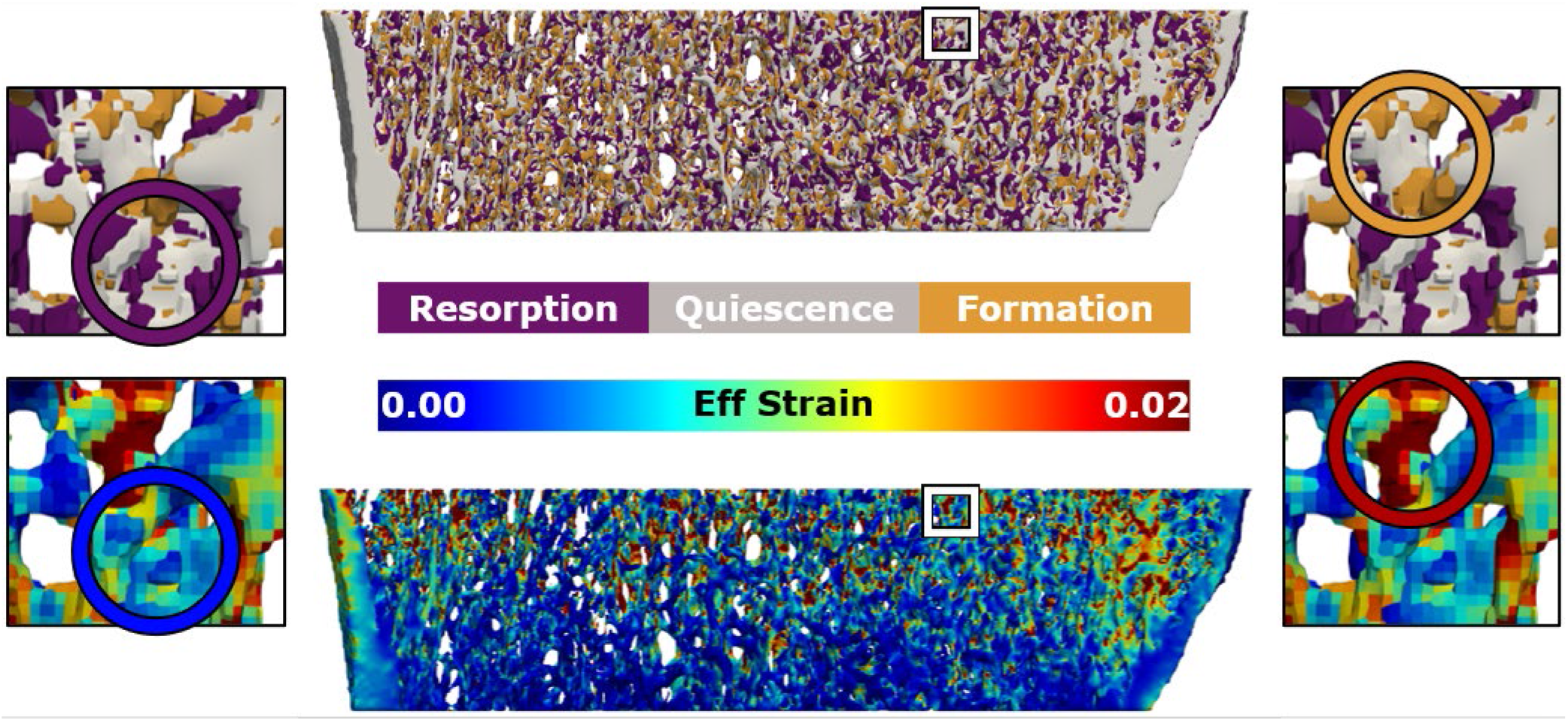
3D reconstruction of remodeling (top) and effective (Eff) strain (bottom) for a representative participant highlighting the prominence of resorption in areas of low effective strain (left) and formation in areas of higher effective strain (right).

## Discussion

While the availability of DXA in the evaluation of patient bone mass and quantity allows for widespread use, the accuracy of measurement is insufficient for use in long-term patient evaluation. This study demonstrated the ability of longitudinal, extended HR-pQCT analysis to identify differences in bone quality and quantity not identifiable at baseline or follow-up using standard clinical analyses (i.e. DXA, static morphometrics, etc.). Among the three groups, altered patterns of remodeling were observed, along with altered mechanoregulative strategies, which were also able to detect differences in dominance of the evaluated arm and sex.

Generally, T-Scores were the lowest for the radius and greatest for the femur. Further, T-Scores of the radius and lumbar spine differed among the three groups, with T-Scores decreasing from NoSupp to OPSupp for both regions, as was expected based on group definitions. Although the femur is often the main target for DXA-based bone quantity scoring, as evidenced by the formulation behind the FRAX calculation for fracture risk^33^, this was the only site that showed no significant differences across groups in the current study. Regarding static morphometrics, differences were observed among groups for Ct.BMD, where LowSupp had the highest value of Ct.BMD and OPSupp the lowest. Since NoSupp had greater radius T-Scores, but lower Ct.BMD than LowSupp, this suggests there may be a compensatory increase in Ct.BMD occurring with initial reductions in areal BMD at the radius. Changes in Tb.Th observed through the duration of the study were affected by group. Interestingly, baseline values of Tb.Th were identical for LowSupp and OPSupp; however, only LowSupp showed significantly lower adjusted values from NoSupp after treatment. In contrast, changes in Ct.Th observed through the duration of the study were affected by sex. Based on the magnitude of change in cortical bone morphometrics, sex appeared to have a greater impact on post-intervention values than group. Previous cross-sectional HR-pQCT studies have revealed sex-based differences in cortical bone morphometrics in both normative and pathologic patient cohorts, with males having consistently higher Ct.Th than females^24,25,34^. When comparing patients with normal bone mass and those with low bone mass and osteoporosis, significant differences in Ct.Th at the radius were detected in female patient cohorts (Normal > Low and OP)^35^, but not in male patient cohorts (Normal = Low and OP)^36^. This would indicate that sex is highly relevant to the morphometric assessment of patients for osteopenia and osteoporosis using the methods outlined herein.

Differences among the three groups were observed for trabecular formation and cortical formation and resorption. Ct.BMD and Ct.Th were predictors for both cortical formation and resorption. LowSupp had the greatest Ct.BMD of the three groups and showed a trend for decreased resorption and formation in the cortical region, while OPSupp had the lowest Ct.BMD and showed a trend for increased formation in both the trabecular and cortical regions. Although no differences in Tb.BMD were observed, both trabecular formation and resorption were predicted by Tb.BMD and BV/TV and trabecular resorption was also predicted by Tb.Th and the time between imaging sessions. In comparison to the other two groups, OPSupp showed a trend for increasing trabecular formation with increasing density, while LowSupp showed a trend for higher trabecular resorption. Combined, these results indicate that both quality and quantity drive formation and resorption volume fractions, with subtle differences in this effect among the three groups. Previous studies evaluating bone remodeling have not evaluated results with respect to morphometrics and densitometrics^30,31,37^, but instead found a relationship between remodeling and age^38^. However, since bone quality and quantity often decrease with age, these previous observations may indirectly support our findings.

Average stiffness values decreased with worsening T-scores, in line with previous studies linking bone loss with a drop in bone mechanical competence^10,35,36,39,40^; however, no significant differences were detected among groups after accounting for pre-intervention stiffness. Further, none of the investigated covariates had a significant influence on post-intervention stiffness. While apparent stiffness appears to be an insensitive parameter to the intervention explored in this study, the interval between baseline and follow-up may have been too short to detect differences in the mechanics at the organ-level. A review assessing the clinical application of HR-pQCT in adult patient populations found less than half of studies assessing bone strength (i.e. stiffness and failure load) reported significant differences between anti-osteoporotic drug treatment and placebo groups, with the majority of trials running for more than 12 months^26^. Of the studies that had a 12 month follow-up interval, only one reported significant changes in response to treatment^41^. Combined with the results of the current work, this indicates the need to establish guidelines for minimum follow intervals in longitudinal HR-pQCT studies.

Few studies have explored changes in *in vivo* strain distribution in longitudinal analysis of human bone, often focusing on regional median or average values^21,31,42^. Although not directly comparable, the patterns of strain distribution in the cortical and trabecular regions from this study are consistent with Johnson and Troy^43^. The bulk of trabecular strains were lower than those in the cortical region; however, peak strains were measured in the trabecular region, likely because of thin individual trabeculae. At the tissue level, group differences were detected in both cortical and trabecular mechanical properties. Within the cortical region, the adjusted strain distribution (10^th^, 25^th^, and 50^th^ percentiles) was lower for LowSupp compared to OPSupp following intervention. Within the trabecular region, group differences were detected in the lowest (5^th^ and 10^th^ percentile) adjusted strain values; however, no significant differences were detected between groups in the post hoc analysis. Although the detected differences were small (3-5%) this could be reflective of small changes in the mineralization at the voxel level that were not detected in the morphological analysis. The material properties assigned for the FE analysis were derived directly from the voxel density via a power law conversion. As such, minute, local increases in density could result in a stiffening of the bone material within the model. Given that the boundary conditions were constant, changes in material stiffness would result in a drop in the measured strain.

Previous imaging studies conducted using lower resolution images have found significantly greater macrostructural and mechanical properties in the dominant radius compared to the non-dominant radius^44,45^. Specifically, bone area and bone mineral content from pQCT-based (330 µm resolution) studies and cortical area and failure load from HR-pQCT-based (82 µm resolution) studies were higher in dominant radii. There, the authors concluded that arm dominance predominantly impacts whole bone structure at a macro level (i.e. total area) and mineralization, but not microstructure. However, image resolution and image quality were lower than that of the current study, thus microstructure likely could not be accurately assessed. More recently, studies using second generation HR-pQCT (62 µm resolution) to study arm dominance have also reported significantly increased macro- and microstructure and mechanical properties in the dominant arm^46,47^. In all cases, increased mechanical stimulus in the dominant arm was considered the main contributing factor to the observed differences. Only one study has reported ambidextrous or equivalent arm dominance, but only one participant identified as such^47^. In the current study, arm dominance had no impact on post-intervention density or morphological parameters. Regarding post-intervention mechanics, ambidextrous arms were found to have significantly higher adjusted strains than the non-dominant (cortical: 10^th^. 25^th^, and 50^th^ percentile strain; trabecular: 75^th^ percentile) and dominant arms (cortical: 25^th^ percentile strain; trabecular: 75^th^ percentile strain). No differences in post-treatment response were detected between dominant and non-dominant arms. Given that the participants in the current study suffered a fracture on the non-investigated arm, the observed differences may have resulted due to changes in the daily loading pattern or usage of the contralateral arm. Participants with dominant arm fractures (non-dominant contralateral arms) likely experienced the greatest overall effect to activity and ability as they may not have been proficient in use of their contralateral arm, while participants with non-dominant arm fractures (dominant contralateral arms) or who were ambidextrous experienced little effect to their activity of daily living. Therefore, patients with ambidextrous arm dominance may have experienced the greatest increase in contralateral arm activity. This increase in stimulus may explain our results; however, as only three participants identified as having ambidextrous arm dominance, this observed effect requires further investigation for confirmation.

Consistent with previous studies which found normal physiological activity levels result in significant relationships between bone formation and FE-derived mechanical stimulus^31,32^, the mechanoregulation analysis revealed strong relationships between local mechanics and remodeling in all groups. Differences were observed in the magnitude of the difference between the resorption and formation threshold values (i.e. width of the lazy zone^31^) as well as the relative position of this zone, between groups and individuals. NoSupp had the widest lazy zone, while LowSupp and OPSupp had narrower lazy zones with thresholds shifted towards lower effective strain values. While these differences were not significant, this suggests patients in LowSupp and OPSupp were more reactive to stimuli (or the lack thereof), requiring less mechanical signal to prompt bone remodeling than for those in NoSupp. Although age was not a significant factor, the two youngest participants in the current study both had higher thresholds for formation and resorption as well as a wider lazy zone than the other participants in NoSupp. Future studies should explore the capabilities of HR-pQCT-based mechanoregulation analysis for addressing differences in participant activity level and age.

This study does have limitations. First, the evaluated participant cohorts were grouped based on a combination of bone density measurements from DXA and prescription of supplements based on blood biomarkers, thus the groups do not allow for direct translation to the effect of either initial bone quality and quantity or supplements, as these factors were not analyzed independently. Further, due to limited availability of patient history, it is unknown the degree to which patients adhered to their prescribed supplements or whether there were potentially relevant clinical factors (fall history, activity level, etc.) that were not included in this evaluation. However, even with a relatively small cohort of patients, this study observed variability in remodeling and mechanoregulation among the groups, which indicates that the sensitivity of HR-pQCT should be further investigated in the clinical evaluation of patients. Second, remodeling and mechanoregulation of the contralateral arm may not be independent of the healing process of the fractured arm. As such, results may have been influenced by the severity of the fracture and change in the dependence on the contralateral arm during healing, as this would vary depending on whether the fractured arm was dominant or not. To address this, dominance of the evaluated arm was included as a factor in our analysis in order to separate this factor from the observed results and was found to affect low to median cortical effective strain.

## Conclusion/Outlook

Longitudinal HR-pQCT was able to detect differences in remodeling and mechanoregulation over periods of 9-12 months, highlighting differences among cohorts of participants with varying bone quality and quantity. Due to the ability of HR-pQCT to detect subtle differences in these participants over time, clinicians should supplement current patient evaluation protocols with microstructural imaging and analysis, such that future diagnosis and treatment strategies, such as supplements for patients with osteopenia and osteoporosis, can be driven by patient-specific bone parameters.

## Methods

A subset of 25 subjects recruited for a time-lapse HR-pQCT imaging study approved by the Ethics Committee of the Medical University of Innsbruck (UN 0374344/4.31) were analyzed (Table 1). Blood samples (35 ml) were analyzed to assess Calcium, Vitamin D3, and Parathyroid Hormone in the Medical University Laboratory, Innsbruck. For patients with low values of the general bone markers, Vitamin D and/or Calcium supplements were recommended and patient compliance was verified. All subjects were above 18 years of age, had a unilateral distal radius fracture, and provided informed consent prior to their participation. To eliminate the convoluting effect of fracture healing on bone remodeling and mechanoregulation, only images of the contralateral, non-fractured radius were analyzed.

### Image Acquisition and Clinical Metrics

Data was obtained as part of an unrelated study investigating fracture healing. HR-pQCT (XtremeCT II, Scanco Medical AG, Brütisellen, Switzerland) images of the contralateral radius were acquired at six time points during the first-year post-fracture (168 slices, 10.2 mm scan length, 60.7 μm isotropic voxels, 63 kV, 1500µA, 46ms integration time, 2304 samples, 900 projections). The standard clinical evaluation was completed at Innsbruck Medical University which provided both densitometric indices, including volumetric bone mineral density (BMD) for the whole bone (Tt.BMD), trabecular (Tb.BMD), and cortical (Ct.BMD) regions, and morphometric indices, including bone volume fraction (BV/TV), trabecular number (Tb.N), trabecular separation (Tb.Sp), mean thickness of the trabecular (Tb.Th) and cortical (Ct.Th) regions, and cortical porosity (Ct.Po) of each study participant. Dual-energy x-ray absorptiometry (DXA) images were acquired three-weeks post fracture for the femur, lumbar spine and radius and were used to quantify the T-score of each subject.

### Dynamic Morphometry

The two images which had the best image quality (lower VGS)^48^. and greatest volume of overlap were used to assess bone formation, resorption, and quiescence. As previously described^30^, the earlier of the two images was transformed using cubic interpolation to be aligned with the imaging coordinate system using the SciPy function library in Python^49,50^. The later of the two images was then rigidly registered and transformed to align with the earlier image using a pyramid-based approach optimized relative to the mean squared error between the two images^51^. Masks of the radius in each image were generated using geodesic active contouring^52^ and used to generate cortical and trabecular masks using the scanner manufacturer’s software.

Images were de-noised with a constrained Gaussian filter (sigma=1.2, truncate=0.8, support=1.0) in Python and thresholds ranging from 200 to 920 mg Hydroxyapatite (HA)/cm^3^ were applied with 120 mg HA/cm^3^ intervals. The two images were then compared to determine voxels that had formed, resorbed, or were quiescent at each mineralized density threshold^53^. Formation and resorption volume fractions were calculated relative to the bone volume at each threshold from the earlier image.

### Computational Mechanics

The overlapping, registered HR-pQCT data were used to generate two micro-FE models for each patient via direct conversion of the image voxels to hexahedral elements (Python 3.7). Scaled, linear elastic material properties, computed directly from the Gaussian filtered (sigma=1.2, truncate=0.8, support=1.0) density data using SciPy^49^, and a Poisson’s ratio of 0.3 were assigned to all elements. High friction compression tests with a prescribed 1% displacement of the total height in the axial direction were performed on all models using 180 CPUs from a CRAY XC40 (Swiss National Supercomputing Centre (CSCS)). Results from models were used to calculate apparent compressive stiffness of the contralateral radii over time and to evaluate longitudinal changes in the effective strain (ε_Eff_) distribution within cortical and trabecular bone. The full-field strain data in each bone compartment was sampled at the 5^th^, 10^th^, 25^th^, 50^th^, and 75^th^ percentiles to enable quantitative comparisons among the NoSupp, LowSupp, and OPSupp groups over the course of the study. Low, median, and relatively high strain percentiles were sampled to better characterize the shape of the strain distributions in lieu of only reporting the median or average ε_Eff_.

Results from the FE analyses were spatially correlated with formation, resorption and quiescent bone volumes to assess local mechanoregulation. Here, conditional probability (CP) curves were generated for the remodeling events identified on the bone surface^54^, connecting the local mechanical environment (ε_Eff_) with the observed formation, resorption or quiescence events. The effective strain distribution for each FE analysis was normalized using the average 99th percentile of the whole cohort and binned at 1% steps for each remodeling event. A group- and bin-wise normalization were used to calculate CP curves for each subject group in accordance with Schulte et al., 2013^54^. A correct classification rate (CCR), measuring the fraction of correctly identified remodeling events using the CP curves^53^, was calculated to summarize mechanoregulation within each group. Additionally, formation (Tf) and resorption (Tr) strain thresholds were derived from the CP curves for each subject at the point for which formation or resorption became dominant, respectively.

### Statistical Analysis

The Python SciPy function library was used to report and evaluate differences in formation and resorption bone volume fractions and mechanics^49^. The average and difference of densitometry and morphometry measures were used for analysis of each group. Group differences were investigated using Kruskal-Wallis one-way analysis of variance (ANOVA) on ranks when data was non-normally distributed.

To determine the effect of treatment on the morphometrics and mechanics of each group, an analysis of covariance (ANCOVA) was performed in R (R version 4.0.4). Here, baseline measurements, group, imaging interval, age, sex, and dominance of the evaluated arm were included as covariates. Time between scans and age were treated as continuous variables, while group (3 levels: NoSupp, LowSupp, OPSupp), sex (2 levels: male, female), and arm (3 levels: dominant, non-dominant, ambidextrous) were categorical variables. Pairwise comparisons were performed for all covariates that had a significant (p<0.05) and near significant (p<0.1) effect on post-intervention values. Tukey’s HSD method was applied to account for multiple comparisons and resulting values are represented as adjusted means ± standard error.

To investigate the parameters which had the largest effect on formation and resorption, partial least squares (PLS) regression was performed on each volume fraction including variables of demographics, group, densitometry and morphometry data, DXA-measured T-scores, and density threshold using the Python Scikit-Learn function library^55^. All variables were scaled and centered prior to analysis. Leave-one-out cross-validation was used to calculate the predictive power of the model and the number of model components was limited to one. The variables were sorted by variable influence on projection (VIP) and the model was run iteratively including additional variables until the Q^2^ score, which is a measure of predictability equivalent to an R^2^ value, no longer improved.

## Data Availability

All data produced in the present study are available upon reasonable request to the authors

## Acknowledgements

The authors acknowledge the Swiss National Science Foundation (320030L_170205), the European Union’s Horizon 2020 research and innovation programme under the Marie Skłodowska-Curie grant (agreement 841316), and the ETH Postdoctoral Fellowship for financial support. This work was supported by a grant from the Swiss National Supercomputing Centre (CSCS) under project ID s841 and s1070.

## Author contributions statement

C.J.C. and P.R.A. conceived the study, developed the methodology, curated the data, performed the analysis, generated visualizations, and drafted the manuscript; C.J.C., P.R.A., and N.O. developed the software; M.B., K.L., and R.M. conceived and supervised the study. All authors reviewed the manuscript.

